# Cancer Risk Assessment Tool: A new general model to estimate cure-rate fraction in patients under tumor therapy

**DOI:** 10.1101/2020.08.27.20183285

**Authors:** Diego C. Nascimento, Pedro L. Ramos, Oilson A. Gonzatto, Gabriel G. Ferreira, Patrícia P.M. de Castro, Renan S. Barbosa, Vinicius O. Boen, Vinicius H. Valentim, Luiz G. Silva, Mariana M. Gomes, Gleice S. C. Perdoná, Francisco Louzada

## Abstract

Cure fraction is not an easy task to be calculated relating probabilistic estimations to an event. For instance, cancer patients may abandon treatment, be cured, or die due to another illness, causing limitations regarding the information about the odds of cancer cure (related to the patient follow-up) and may mislead the researcher’s inference. In this paper, we overcame this limitation and proposed a risk assessment tool related to the lifetime of cancer patients to survival functions to help medical decision-making. Moreover, we proposed a new machine learning algorithm, so-called long-term generalized weighted Lindley (LGWL) distribution, solving the inferential limitation caused by the censored information. Regarding the robustness of this distribution, some mathematical properties are shown and inferential procedures discussed, under the maximum likelihood estimators’ perspective. Empirical results used TCGA lung cancer data (but not limited to this cancer type) showing the competitiveness of the proposed distribution to the medical field. The cure-rate is dynamic but quantifiable. For instance, after 14 years of development/spread of lung cancer, the group of patients under the age of 70 had a cure fraction of 32%, while the group of elderly patients presented a cure fraction of 22%, whereas those estimations using the traditional (long-term) Weibull distribution is 31% and 17%. The LGWL returned closer curves to the empirical distribution, then were better adjusted to the adopted data, elucidating the importance of cure-rate fraction in survival models.

## 1. Introduction

Cancer is a disease that inflicts a significant portion of the world population. According to Cancer Research UK, there were 17 million new cancer cases in the year 2018 and 9.6 million deaths caused by cancer in the same year [1]. Lung cancer alone is responsible for the most significant number of deaths (1.8 million deaths, 18% of the total), according to the World Health Organization [14]. There has been a tremendous governmental and research institutes effort to minimize the causes of cancer and understand the efficiency of the treatments for the disease. It results in a significant amount of resources spent in order to improve people’s quality of life.

An important issue that usually appears during the treatment of the disease is related to the proportion of patients cured. Usually, correctly estimating the cure fraction of the patients is not an easy task. During the experiment, different reasons may cause the researcher to lose patient follow-up, such as abandonment of the treatment, a patient may be cured or die due to another illness, to list a few. These patients are usually considered as censored information. However, such information plays a vital role during the correct estimation of cured patients. Analyzing these situations, we can find the long-term survival models which take into account a proportion of patients that may not die due to the event of interest.

Berkson and Gage [3] introduced a critical model that considers the existence of two subgroups in the population. The first is susceptible to the event of interest, while the second is non-susceptible (cured). This model is usually referred to as a standard mixture model. Chen et al. [5] discussed the promotion of a long-term survival model, assuming that the latent activation mechanism that leads to the chances of an individual becoming susceptible or not follows a Poisson distribution. Recently, many new long-term distributions have been proposed (see, for instance, Rodrigues et al. [17] and the references therein). The distributions cited above are general families and depend on the choice of a baseline model. Cordeiro et al. [7] considered the Birnbaum-Saunders distribution as a baseline when the general family is obtained from a negative binomial distribution. Ramos et al. [16] considered the Fréchet distribution in the standard mixture model. Further, Ramos and Louzada [12] derived a long-term weighted Lindley distribution based on the distribution introduced by Ghitany et al. [9].

In this article, we introduced a new cure rate model, named long-term generalized weighted Lindley (LGWL) distribution, to describe and estimate the cure fraction in a study of lung cancer, as summarized in Figure 1. This distribution considers as a baseline the generalized weighted Lindley distribution (Ramos and Louzada [12]), a critical model that unified different generalizations of the Lindley distributions such as the weighted Lindley [9] and power Lindley [8]. For the proposed model, essential mathematical functions are presented. The inferential procedure is conducted under the maximum likelihood estimators. The likelihood equations are derived and can be used to achieve the estimates of the parameters. A simulation study is presented with synthetic data, which shows that our estimation approach can be used satisfactory to obtain the estimates in real problems. This new distribution, LGWL, can be used to describe the life expectancy of patients with lung cancer.

**Figure 1.**
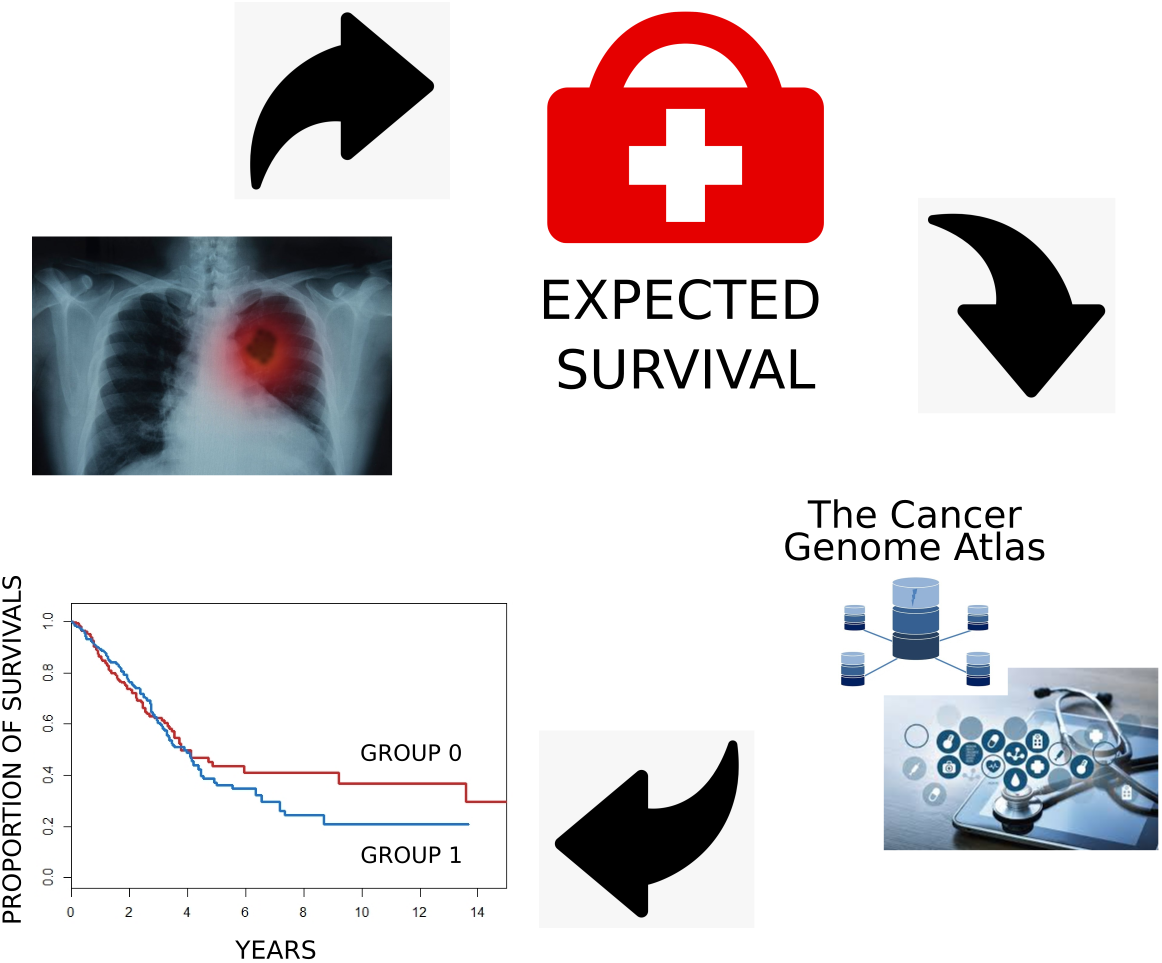
Visual summarization of this article. The process of quantifying the risk starts on cancer detection, then estimating its survival rate, in which was based on the cancer genome atlas, by considering a cure-rate fraction for two different groups.

Additionally, the class of long-term models was considered because it estimates the cure fraction in the data presenting long-term survivors (immune or cured). For instance, the set extracted from [11] using the public data set from The Cancer Genome Atlas (TCGA) set. TCGA is a program that has been generating for the last 12 years over 2.5 petabytes of data related to cancer under researchers’ efforts in different fields supported by the National Cancer Institute and the National Human Genome Research Institute. In this work, we focus on cancer lung motivated by the complexity of the many subtypes defined by this pathology [6]. However, other types of cancer data can be obtained from the cited study. The data set initially contained 629 patients with lung cancer with eight variables that are most relevant to describe the cure of this pathology.

The remainder of the paper is organized as follows. In section 2, there is detailed information about the long-term generalized weighted Lindley distribution and its mathematical properties. Section 3 presents the motivation and mathematical properties of the long-term survival models. Section 4 contains a simulation study to evaluate the performance of the estimation method that was selected. Sections 5 and 6 show the application of the model on real data and some of the results. Finally, Section 7 summarizes the study. Appendix A contains a detailed overview of the estimation process and methods of the distribution parameters.

## 2. The generalized weighted Lindley distribution

Ramos and Louzada [13] recently proposed the generalized weighted Lindley (GWL) distribution, which can be used to describe a wide range of problems. Let T be a random variable with a GWL distribution, then its probability density function (PDF) is given by

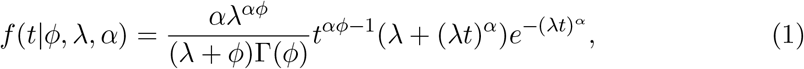

for all *t >* 0, where 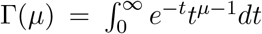 is the gamma function, *ϕ >* 0, *α >* 0 and *λ >* 0 are the shape, scale and shape/scale parameters, respectively. One of its peculiarities is that the hazard function can have an increasing (*ϕ* ≥ 1) or bathtub (0 *<ϕ<* 1) shape.

The mean and variance related to the GWL distribution have closed-form expression which is given by

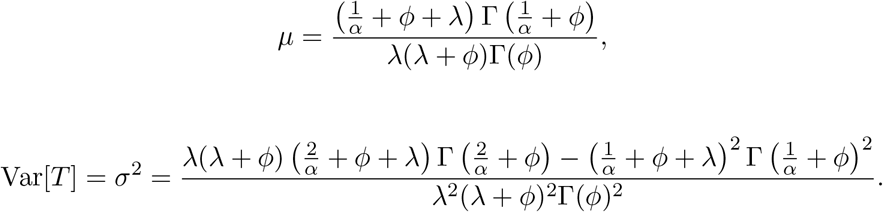

Two important functions related to the GWL distribution are the survival and hazard rate functions. The former returns the probability of a patient’s survival beyond a specified time. On the other hand, the hazard function measures the instantaneous risk of death, given that the patient is still alive. These functions are given by

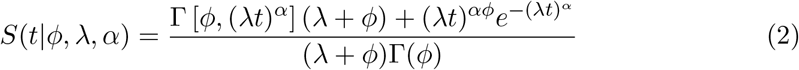

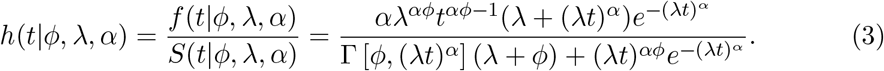

where 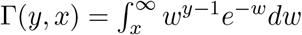 is the upper incomplete gamma function.

The behavior of the hazard function (3) when *t* → 0 and *t* →∞ are, respectively, given by:

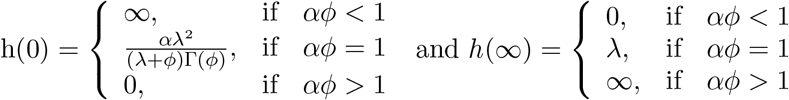

The principal authors have proved that the hazard function *h*(*t*) of the GWL distribution can have different shapes such as increasing, decreasing, bathtub, unimodal, or decreasing-increasing-decreasing shape. This property makes GWL distribution a flexible model to describe lifetime data. In this sense, we consider the GWL a baseline model to construct a new long-term survival model.

## 3. Long-term survival Model

In survival analysis theory, it is commonly assumed that the event of interest will happen. However, there are many cases where the event of interest does not occur. For instance, in cancer therapy, the patient may be cured after the treatment and may not die due to cancer remission. In these situations, the patient cannot be considered to have achieved the event of interest. The usual lifetime models cannot be used to describe such situations. This problem is overcome by including a cure fraction in the modeling. Such cure fraction represents the patients that did not die due to the event of interest. These models are called long-term survival models, in which the population is divided between immune individuals (insusceptible) and non-immune (susceptible). Thus, immune individuals will certainly be censored during the experiment, while susceptible individuals will fail (or die). Hereafter, *π* is considered the parameter associated with the cure fraction, while (1 − *π*) is related to the proportion of the non-immune group. Berkson and Cage [3] have proposed the standard mixture cure model given by

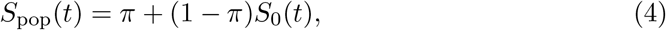

where *π* ∈ (0, 1) and *S*_0_(*t*) denotes the survival function related to the susceptible group. The obtained survival function (4) is improper and expresses the survival function related to the whole population. Note that as time goes to infinity, the limit tends to *π*, i.e., the proportion of cure in the population.

From the survival function (4), the probability sub-density function (PSDF) is easily obtained

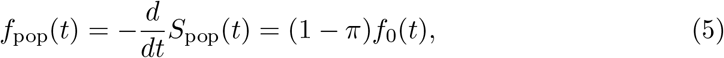

where *f*_0_(*t*) is the baseline PDF for the non-immune individuals. The hazard function can also be given by

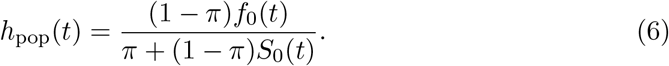

Under the assumption that *f*_0_(*t*) follows a GWL distribution, the PSDF of the long-term generalized weighted Lindley (LGWL) distribution is given by

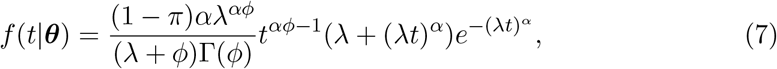

where ***θ*** =(*ϕ, λ, α, π*) denotes the parameter vector. By considering (2) and (4), the improper survival function of the LGWL distribution is obtained:

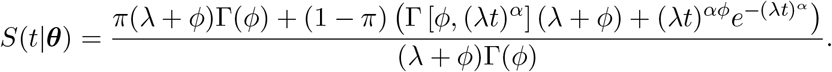

Figure 2 shows the shape of the PSDF and the improper survival function of the LGWL distribution, for some parameter values.

Finally, the hazard rate function of the LGWL distribution is given by

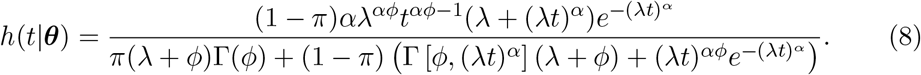

Figure 3 shows the shape of the hazard function of the LGWL distribution.

**Figure 2.**
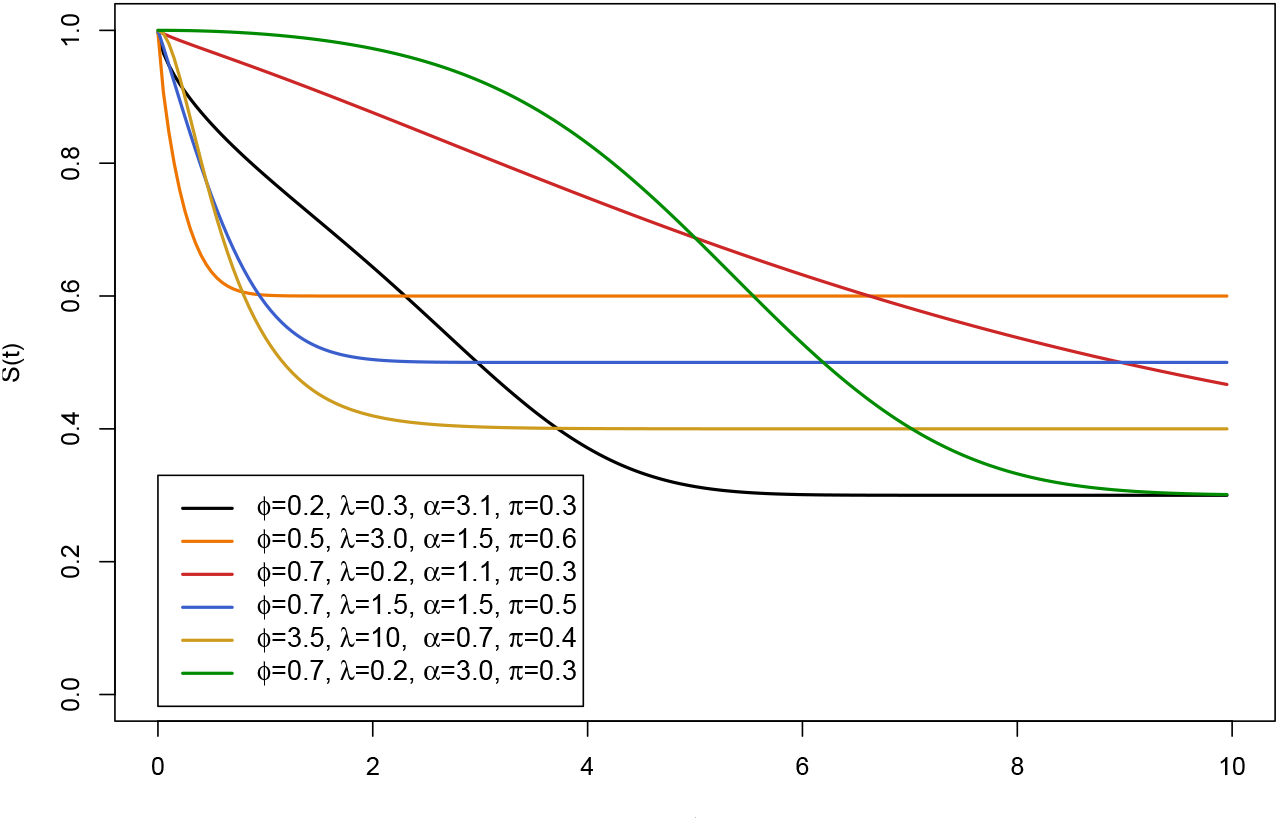
Survival shape of the LGWL distribution for different values of *ϕ*, λ, α and π.

**Figure 3.**
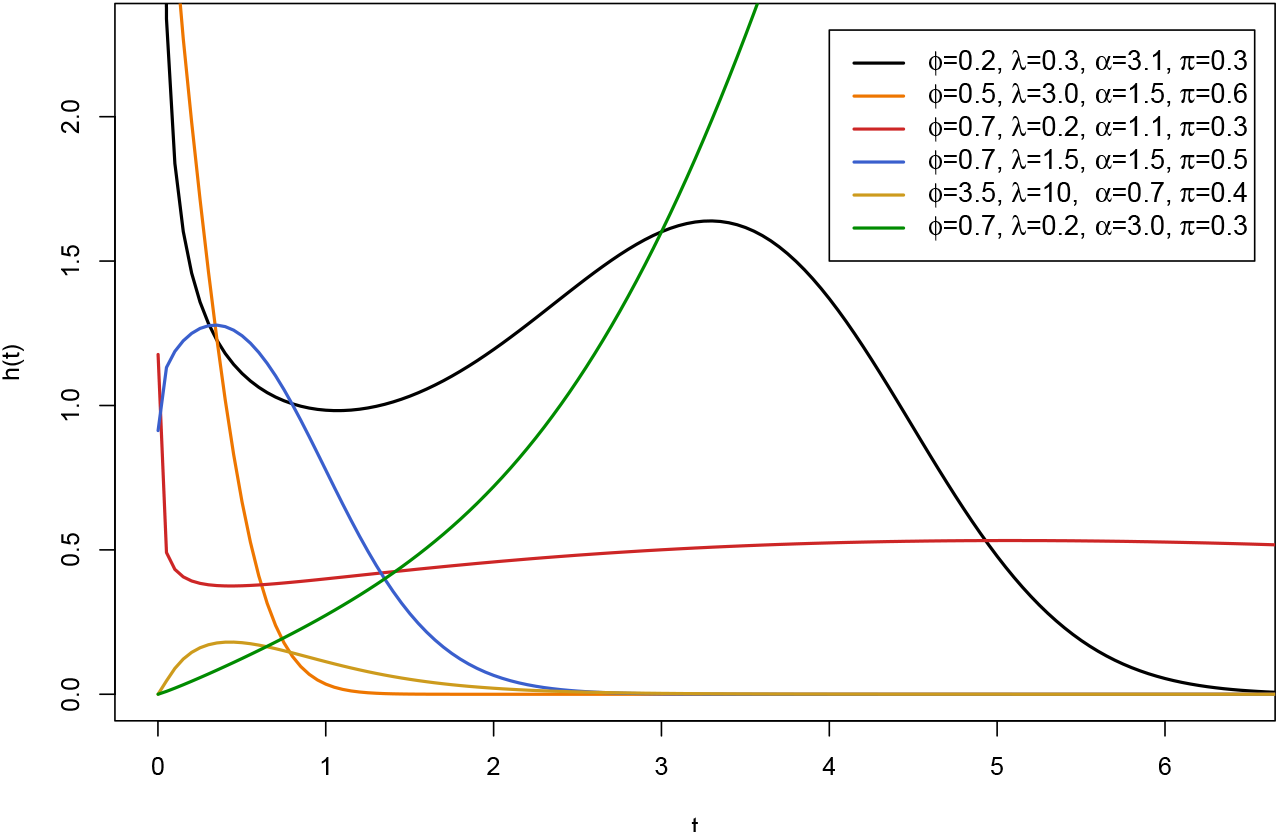
Hazard shape of the LGWL distribution for different values of *ϕ*, λ, α and π.

## 4. Simulation Study

Monte Carlo simulation technique is used to evaluate the performance of the maximum likelihood estimate (MLE) method. The simulation is done by computing the mean relative estimate (MRE) and the mean square error (MSE), which is given by:

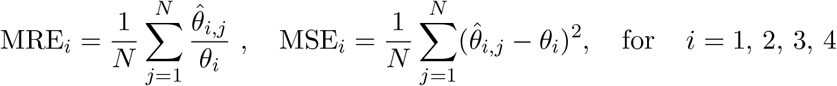

where N = 10,000 is the number of simulations performed. In other words, N represents the estimated amount of ML. Covered probabilities (CPs) of the 95% asymptotic confidence intervals were also computed. Assuming the chosen values for the simulation parameters, it is expected that MLEs will generate MRE values close to one with small MSE values. Besides, it was expected that the proportion of intervals containing the true value of the parameter would be close to 0.95, as a 95% confidence interval was considered.

The simulation was performed using the statistical software R [15] and the package used to maximize the log-likelihood function (9) was the maxLik. The following parameter values were considered to perform the simulation: *ϕ* =0.8, *λ* =0.5, *α* =1.5 and *π* =0.4 (Figure 4); *ϕ* =1.2, *λ* =0.5, *α* =0.97 and *π* =0.25 (Figure 5), where n = {50, 60*,…*, 300}. For this simulation, the assumed censoring scheme was random, and two different scenarios were considered, where the long-term survivals fractions were 0.4 and 0.25, respectively. The simulation was also applied considering new values for the parameters and similar results were obtained. In addition, the procedure converged and ended without multiple maxima.

**Figure 4.**
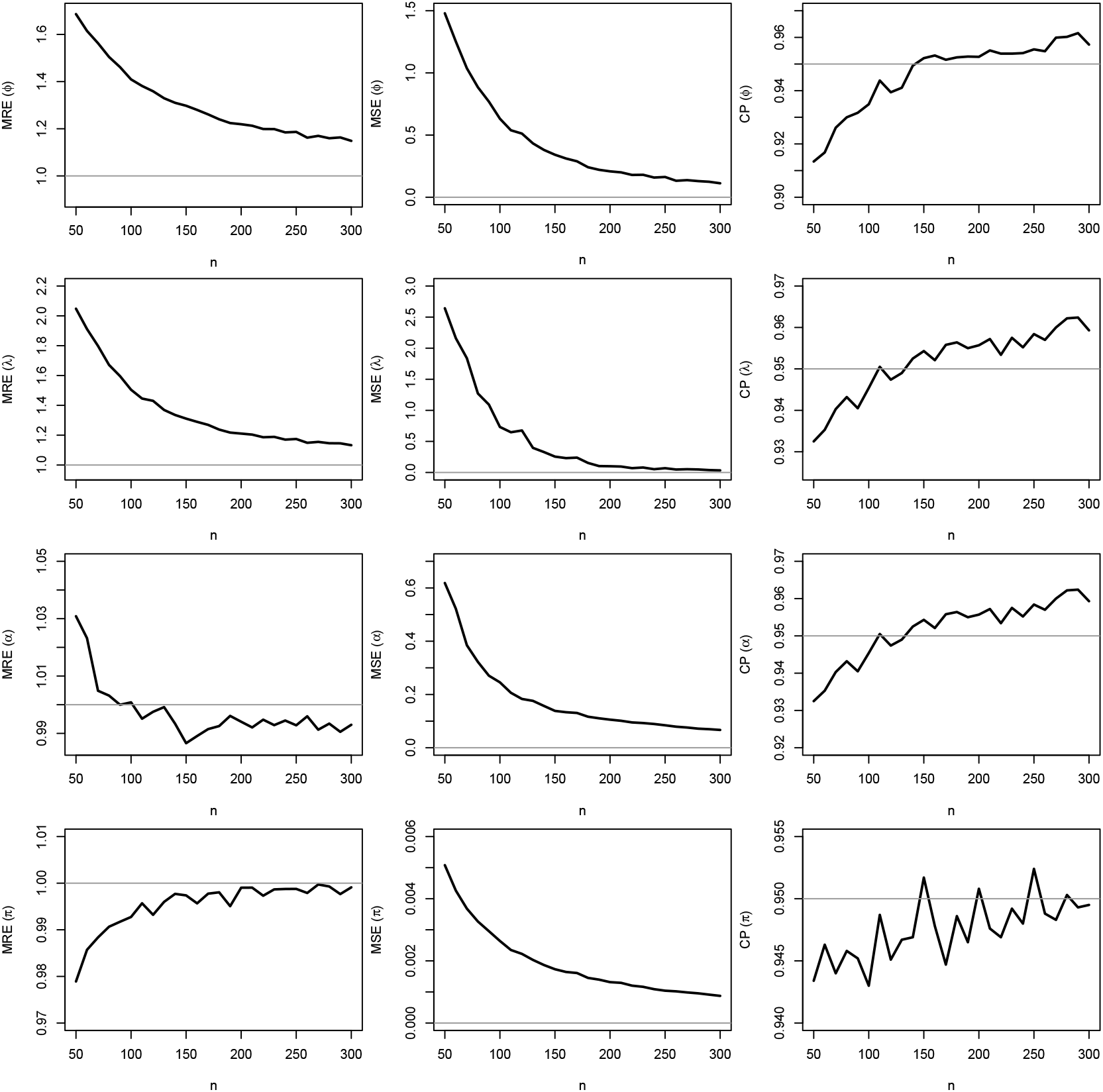
MREs, MSEs and CPs related to the ML estimates of *ϕ* =0.8, λ =0.5, α =1.5 and π =0.4, for N = 10, 000 simulated samples and different values of n.

**Figure 5.**
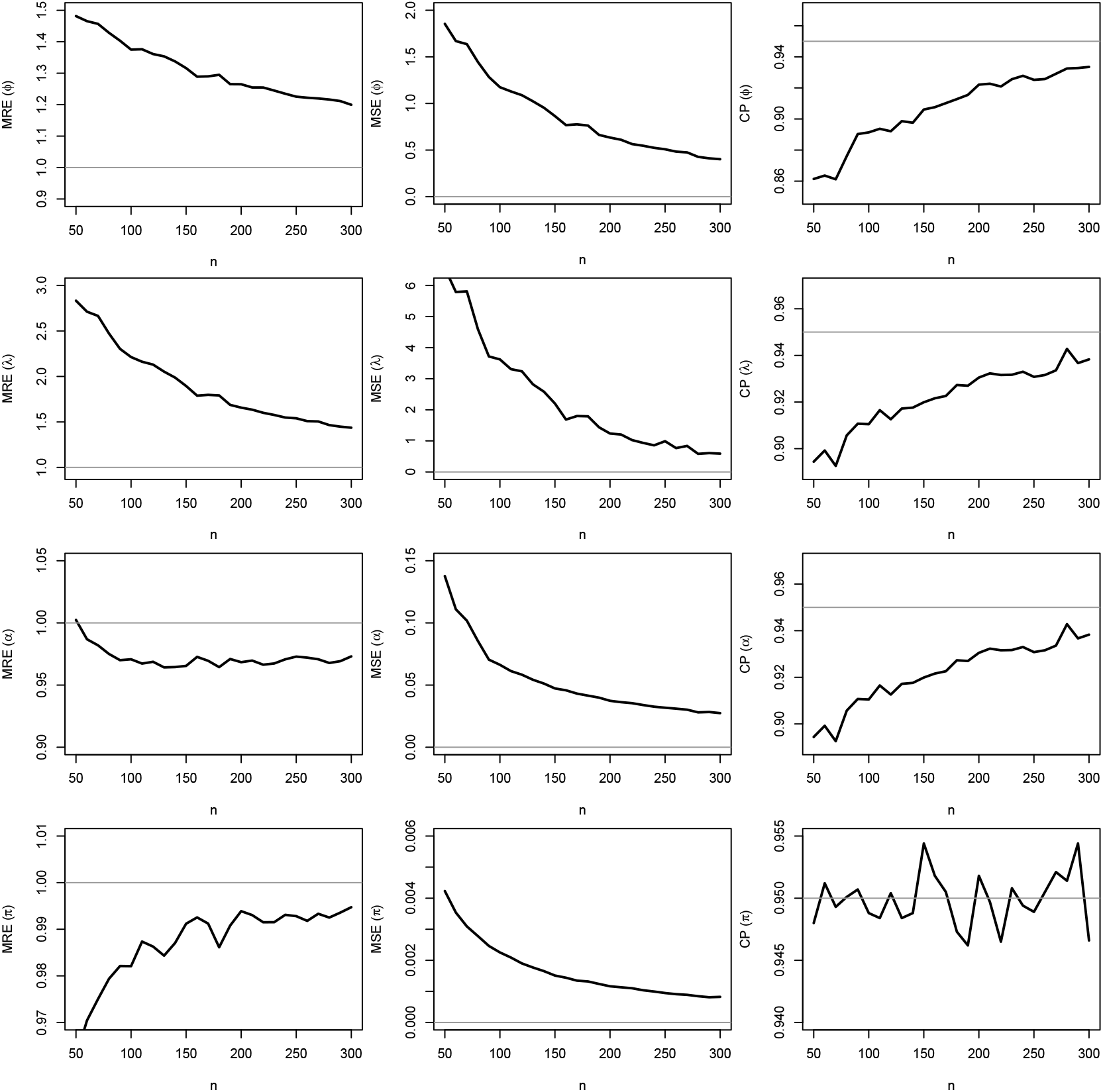
MREs, MSEs and CPs related to the ML estimates of *ϕ* =1.2,λ =0.5, α =0.97 and π =0.25, for N = 10, 000 simulated samples and different values of n.

Figures 4 - 5 bring the results of MREs, MSEs and CPs of the ML estimates of *ϕ*, *λ*, *α* and *π* for each value of *n* presented before. The horizontal line shown in the plots corresponds to MRE, MSE, and CP values equal to one, zero, and 0.95, respectively. As it can be noted from the plots, MRE values tend to one, and MSEs approach to zero as *n* grows, meaning that the MLEs of *ϕ*, *λ*, *α* and *π* are asymptotically unbiased.

Besides, as *n* becomes large, CPs tend to 0.95, which indicates that good coverage properties may be deliberated for the MLEs. More importantly, the results are very close to the true ones specially for samples greater than 150, showing the convergence accuracy of the proposed estimator. Thus, the ML estimation method will be relevant in practical applications, as shown in the next section.

The present study can also be generalized by considering covariates in long-term survival to improve the prediction. For instance, it is possible to use the logistic link function given by

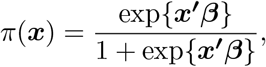

where ***β*** =(*β*_0_*, β*_1_*,…,β_p_*)′ is the vector of parameters related to the vector of covariates ***x*** = (1*, x*_1_*,…,x_p_*).

## 5. The Cancer Genome Atlas Data

A sample from the Cancer Genome Atlas data (also known as TCGA) is considered in the present work. The TCGA set came from a 10-year project, and it can be accessed at GDC Data Portal, the original data contains more than 11,000 patients with 33 different types of cancer (where the top 3 are breast invasive carcinoma, kidney clear cell carcinoma, and lung adenocarcinoma). However, the focus herein is to analyze lung adenocarcinoma (a subtype of non-small cell lung cancer), which is related to the lifetime (in years) of 629 patients (5.72% of the set). Figure 6 shows the empirical distribution of the top 3 types of cancer in the data set, including non-small cell lung cancer.

**Figure 6.**
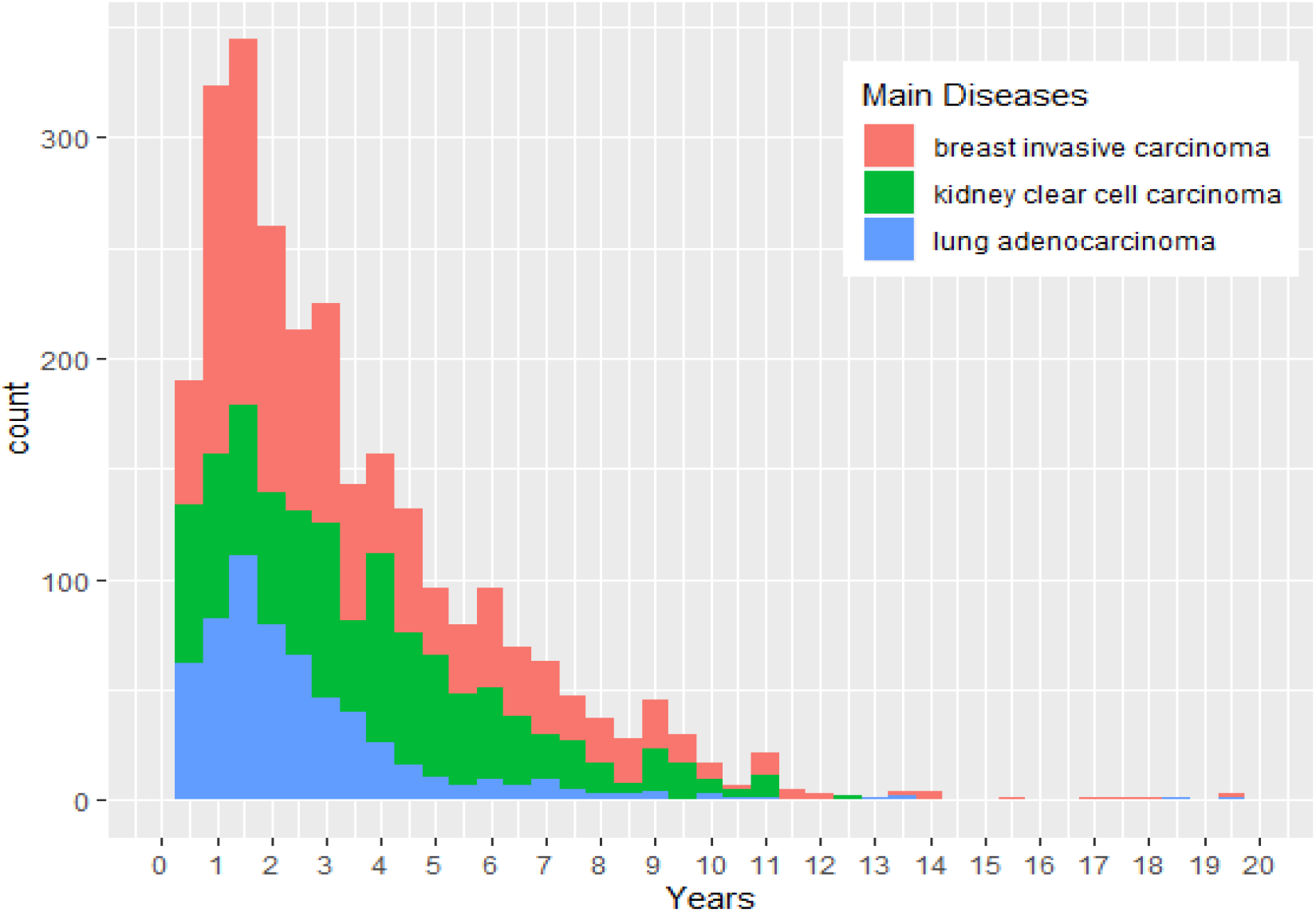
Absolute frequency (in years) of death related to the top three types of cancer. The non-small cell lung cancer in the TCGA set is shown in blue.

Elements related to the cancer survival rate are related to the efficiency and survival benefits on patients treated with the available therapeutic approaches [4]. The variables associated with the patients’ lifetime can be viewed as detailed in Table 5.

**Table 1.** Description of all relevant variables in the TCGA set.

| Variable | Description |
| --- | --- |
| Event | Event, in this case is the overall survival event |
| OS | Overall survival time |
| Age at initial pathological diagnosis | Age of the person when the first diagnosis was established |

A summary of the dataset is also presented, containing metrics for the numeric variables, such as mean, median, min, max, and the number of levels for the categorical ones in Table 5.

**Table 2.** Summary for the numeric variables of the TCGA set

| Name | NA | Mean | SD | Median | MIN | MAX |
| --- | --- | --- | --- | --- | --- | --- |
| OS | 0 | 930.7 | 892.0 | 697 | 0 | 7248 |
| Age | 0 | 65.38 | 9.9 | 66 | 33 | 88 |

Xiong et al. [19] addressed the issue of the data-driven model towards observed survival interval, defining the period from TCGA sampling to patient death or last follow-up as the time interval. Moreover, some features such as smoking and age were considered to gather more information towards the cure-rate association in cancer patients [18]. In Table 5, it is observed that the mean of overall survival time (in days) is 930,779. However, the standard deviation is 892,036, which shows a high variability of the overall survival time. The mean and median of age indicate that the majority of the patients are middle-aged.

**Table 3.** Summary for categorical variables of the TCGA dataset

| Categorical Data Summary |  |  |  |  |
| --- | --- | --- | --- | --- |
| Name | Type | NA | disp | nlevs |
| Event | Factor | 0 | 0.28 | 2 |

Araujo et al. [2] discussed the importance of early diagnosis in lung cancer, and its survival rates, highlighting the time needed to diagnose the illness and the beginning of the palliative treatment. Assuming the patient survival as a variable named *Event*, it is evident that in the majority of cases, patients succeed on surviving in lung adenocarcinoma therapy, indicated by class 0 (zero), compared to the non-survivors represented as class 1 (one), shown in Table 5. Moreover, this is still a low survival rate compared to other types of cancer and modeled as follows.

**Table 4.** Frequency of the variable event

| Event | Frequency | Percentile Freq. |
| --- | --- | --- |
| Person Survived (0) | 389 | 0.6184 |
| Person did not Survive (1) | 240 | 0.3816 |

In Figure 7, two groups of events were analyzed, taking into consideration the age of the patients, and the LGWL distribution was compared with other models, such as Kaplan-Meier (KM), Long-term Weibull (LW) and Long-term weighted Lindley (LWL) distribution. It is possible to note that, at first, the empirical data set intersects, which indicates that the hazard functions are not proportional. In this case, the Cox proportional-hazard model is not adequate [10]. When the test of proportionality was applied, it returned a p-value of 0.025, indicating a violation of the proportionality assumption. From the adjusted parametric models, it was observed that the LW and LWL are not so close to the empirical model, while the LGWL returned closer curves of the KM.

**Figure 7.**
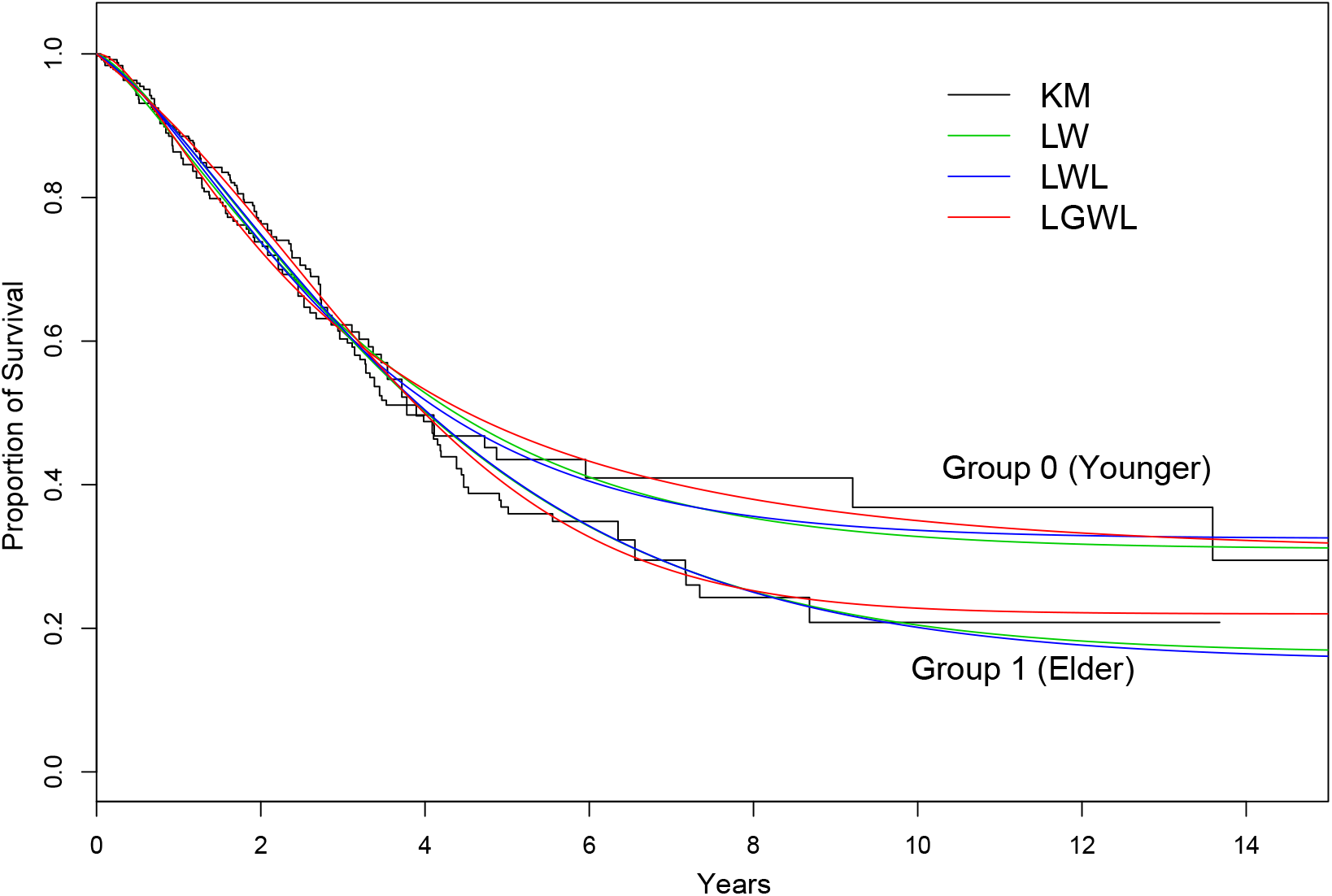
Event counts by cancer type separated hin two groups based on age. The solid black line represents the Kaplan-Maier (KM) empirical distribution. The green line represents the Long-term Weibull (LW) distribution, blue the Long-term weighted Lindley (LWL), and red line the long-term generalized weighted Lindley (LGWL). The LGWL returned closer curves to the empirical distribution, then better adjusted to the presented data.

Table 5 presents the Akaike information criterion (AIC) values for different long-term lifetime models using Groups 0 and 1. It can be observed from this table that, although subtle, the LGWL distribution provides a better fit to these data since the adjusted distribution has the lowest value. An essential result of the present work is that we were able to observe such unimodal hazard behavior while the current long-term Weibull distribution cannot fit this type of data. Hence, the new model is an efficient tool to estimate the cure fraction in a clinical trial.

**Table 5.** AIC values for the fitted distributions for group 0 (younger patients) and group 1 (elder patients).

|  | Weibull | LW | LWL | LGWL |
| --- | --- | --- | --- | --- |
| Group 0 | 547.80 | 533.10 | 533.20 | 531.37 |
| Group 1 | 840.52 | 838.90 | 837.40 | 836.77 |

The parameter estimates of the LGWL distribution were obtained using the same procedure as described in Appendix A. The patients were divided into two age groups to determine if their age influences on the survival time. This covariable was created to check if there are significant differences in survival time between patients who had cancer up to 69 and those who had the disease at 70 years of age and more. Patients up to 69 years old were classified as “age = 0,” and those aged 70 and older were labeled “age = 1”. Table 6 exhibits the ML estimates, the standard errors (SE) and 95% confidence intervals (95% CI) for ages 0 and 1.

**Table 6.** ML estimates, SE and 95% CI for the parameters of the LGWL distribution, considering the TCGA set for ages 0 and 1

|  | Group 0 (Younger) |  |  | Group 1 (Elder) |  |  |
| --- | --- | --- | --- | --- | --- | --- |
| Parameter | $\hat{\theta}$ | SE | 95%CI | $\hat{\theta}$ | SE | 95%CI |
| $\hat{\phi}$ | 5.902 | 0.1672 | (5.101 ; 6.703) | 0.647 | 0.0304 | (0.304 ; 0.988) |
| $\hat{\lambda}$ | 23.208 | 3.8835 | (19.346 ; 27.070) | 0.316 | 0.0023 | (0.221 ; 0.410) |
| $\hat{\alpha}$ | 0.427 | 0.0008 | (0.370 ; 0.484) | 1.491 | 0.0878 | (0.910 ; 2.072) |
| $\hat{\pi}$ | 0.304 | 0.0058 | (0.154 ; 0.454) | 0.220 | 0.0027 | (0.117 ; 0.322) |

As can be seen in the previous table, the results show that it is possible to estimate the cure fraction (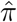) related to each group. Since the cure fraction was different for each age group, it is possible to assume that as the patient’s age progresses, the cure-rate fraction increases, meaning that younger patients have a higher probability of surviving lung cancer than older patients.

The measure of centrality of the hazard distribution (*α*) indicates some centrality of the event rate at time t conditional on survival until time t or later (e.g., the centrality force of mortality). Additionally, from the estimated parameter values, we observe that the hazard rate has unimodal behaviors: for the younger group, the most dangerous period is close to 1.3 years after the pathological diagnosis. For the older group, the higher risk period is close to 4 years after the diagnosis.

## 6. Interactive Web App

Thus, adjusting some survival models, considering the long-term, an interactive online tool was implemented to unravel the gain of the proposed model, by visualizing the applicability of this class of models in the medical field. The URL https://cemeai.shinyapps.io/cancer_tool provides this tool to explore better the potential of some survival methods with the cure-rate fraction.

The developed framework has mainly three elements: i) the survival model selection, ii) cards displaying information about the estimated cure fraction, and iii) three graphical reports regarding the adjusted survival curve, the instant risk and the risk ratio between the groups under study, as shown in Figure 8. The central graph (left-hand) shows the selected model(s) fit the Kaplan-Meier non-parametric estimator. Moreover, it is possible to indicate the quality of the model’s adjustment through obtaining point estimates, iteratively, for the survival of the groups (young and elderly patients) over time. The adjusted models can also be evaluated through their instantaneous hazard functions (right-hand top chart), and time-varying odds rates (right-hand bottom chart) of the hazard of the groups.

**Figure 8.**
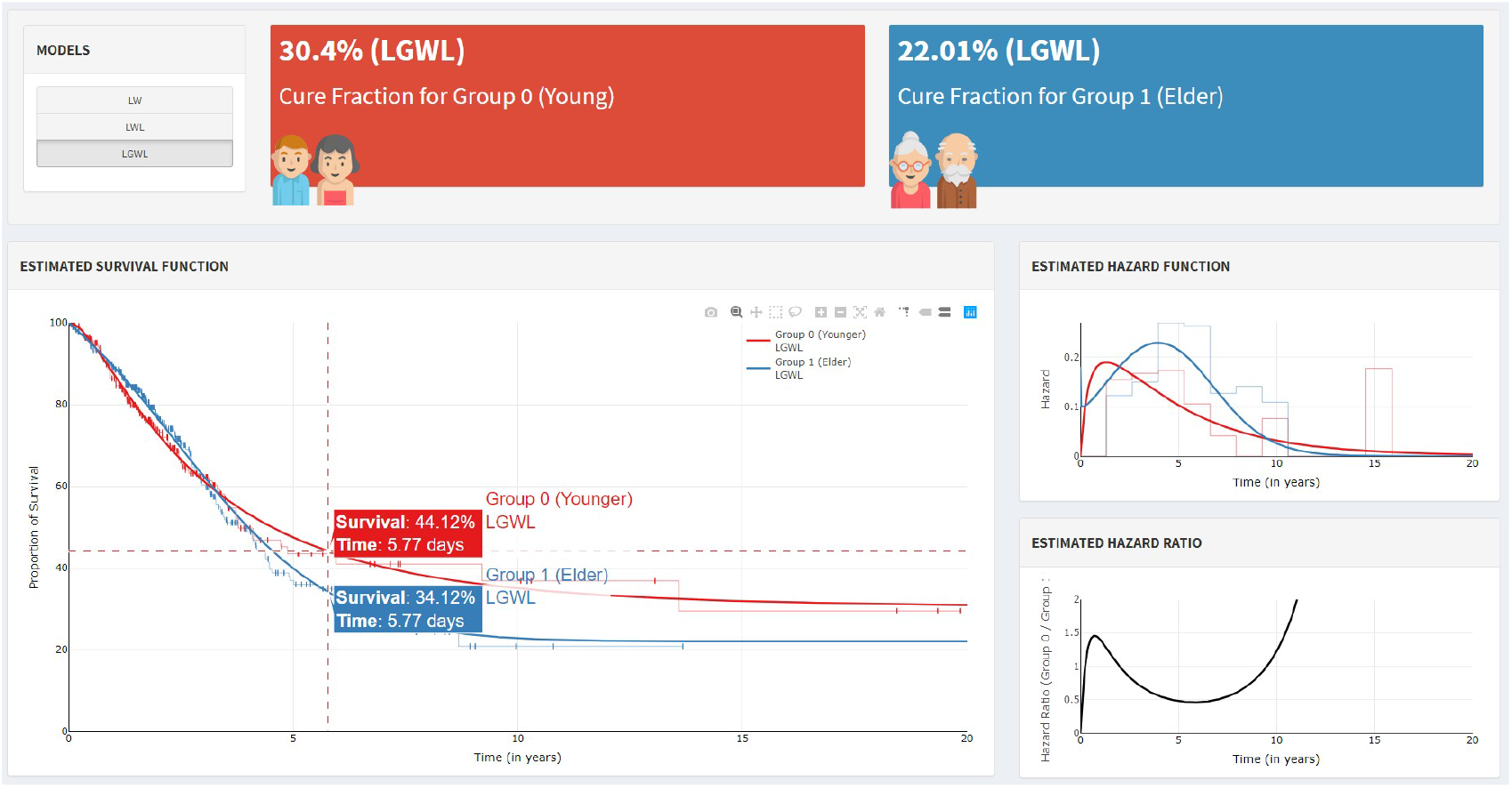
Cancer Risk Assessment Framework, adopting the cure-rate fraction general models. On the Top left, the user can select the models, and the top center and right shows the estimated cure-rate fraction for both groups (younger vs. elder). The left-hand chart represents the adjustment of the selected model based on the empirical function (Kaplan-Meier). The right-hand top chart presents the instantaneous failure rate and Right-hand bottom chart the odds rate among the groups.

In the right-hand graphs, we can see the estimated risk’s general behavior based on the selected model. The instantaneous hazard risk function can provide an overview, among time, when the risk is maximum, or when it is growing (or decreasing), for each group. On the other hand, the graph-related hazard odds ratio shows a relative comparison of each group’s risks under study, indicating the moment and the magnitude when the risk of one category becomes higher (or lower) than the other. For instance, for a fraction of years, in early detection, the younger group cares a higher risk than elders (right-hand top chart in Figure 8).

## 7. Discussion

In this article, we proposed an essential extension of the generalized weighted Lindley. This distribution takes into account individuals who are long-term survivors of lung cancer (event of interest). The Long-term generalized weighted Lindley (LGWL) distribution can estimate the cure-rate fraction of the patients, even considering the posterior period of the study (named long-term). We discussed different mathematical properties for the proposed model, such as the mean, variance, survival function, and hazard rate function.

The lifetime of patients that had lung cancer, supported by the database from the National Cancer Institute and the National Human Genome Research Institute, show empirical survival functions for different types of age. These results imply different patterns regarding the cure fraction for two main groups (group 0 is composed of the patients who had lung cancer before the age of 70, and group 1 is composed of the patients who are 70 years of age and older).

Moreover, by using the LGWL distribution, medical doctors will have a reference for survival estimation based on the development and spread of cancer as an inverse problem. While the patient has the disease, a different survival rate must be associated with each period, since the patient’s chances of survival should decrease over the years. For instance, in the second year of the development of lung cancer, both groups of patients (younger and older age) present an 80% cure-rate fraction. However, in the tenth year, the group of patients who are younger than 70 presents a 40% rate, whereas the group of elders presents a 20% cure fraction (according to Figure 7).

Based on the estimated cure fractions, after 14 years of the development/spread of lung cancer, the group of patients under the age of 70 had a cure-rate fraction of 0.322 (which means that around 32% of these patients will survive), while the group of elderly patients presented a rate of 0.2204 or 22%, whereas those estimations using the long-term Weibull distribution is 31% and 17%, respectively (underestimating the cure-rate fraction of the elder group).

## Data Availability

.

## Acknowledgements

Pedro L. Ramos acknowledge the support from the São Paulo State Research Foundation (FAPESP process 2017/25971-0). Francisco Louzada acknowledges support from the S ao Paulo State Research Foundation (FAPESP Processes 2013/07375-0) and CNPq (grant no. 301976/2017-1). Gleici S.C. Perdoná acknowledge the support from Fundação de Apoio ao Ensino, Pesquisa e Assistência do Hospital das Clínicas da Faculdade de Medicina de Ribeirão Preto da Universidade de São Paulo (FAEPA) and to FAPESP.

## Appendix A

In this appendix, we discuss the necessary steps to conduct the inference in the parameters of the proposed model. Suppose that the lifetime of the *i*-th patient may not be observed and it is subject to right-censoring. Moreover, consider that the random censoring times *C_i_*’s are independent of the true lifetimes *T_i_*’s, and the distribution of *C_i_*’s does not depend on the parameters governing the distribution of *T_i_*’s. Then, for a sample of size *n*, the dataset is 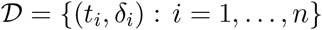, where *t_i_* = min {*T_i_, C_i_*} and *δ_i_* = *I*(*T_i_* ≤ *C_i_*), with *I*(·) denoting an indicator function. This random censoring scheme has as special cases the types I and II censoring schemes. The maximum likelihood method is the most widely used approach for estimating parameters, since it provides estimators (MLEs) that have several desirable properties, such as asymptotic efficiency, consistency and invariance. The likelihood function is given by

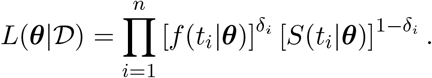

Let *T*_1_*,…,T_n_* be a random sample of size *n* from the LGWL distribution (7). Then, the likelihood function, considering data with random censoring, is given by

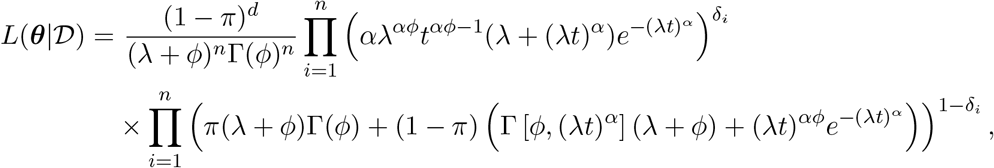

where 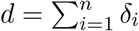 is the number of censoring in the sample.

The log-likelihood function is given by

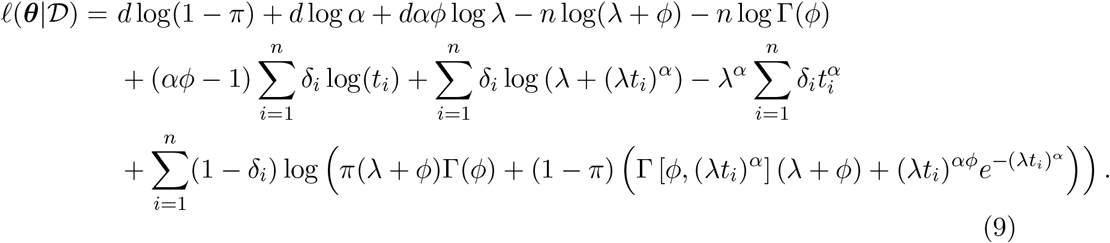

The MLEs 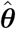 of *θ* are obtained from the solution of 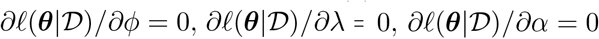 and 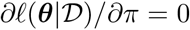. Hence, the likelihood equations are given as follows:

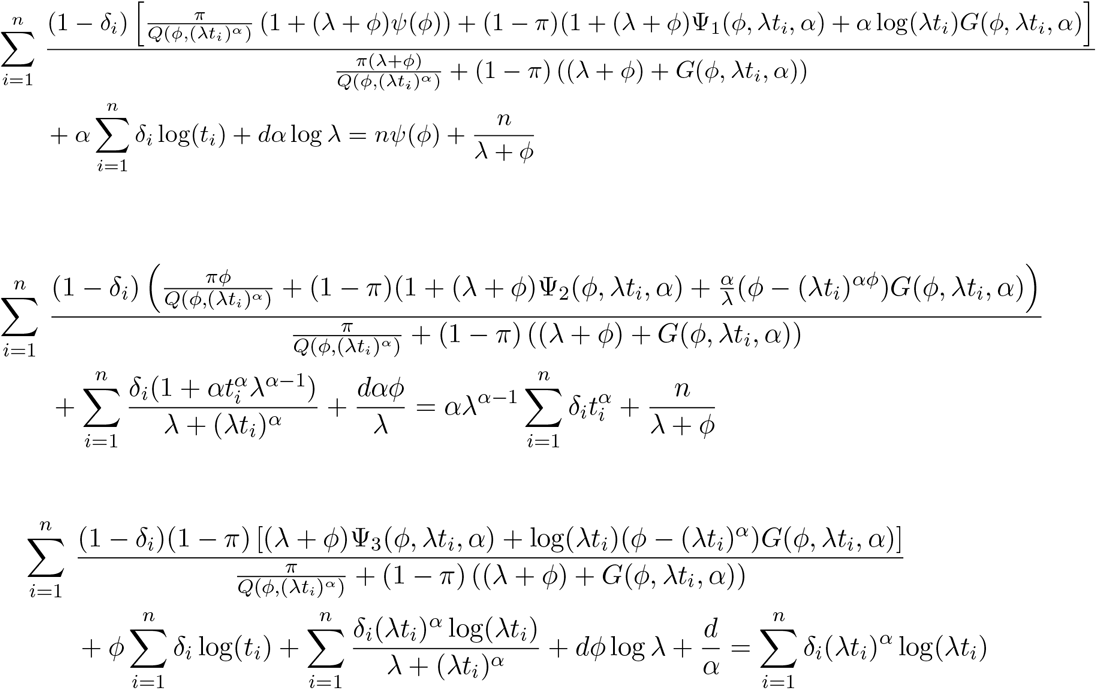

and

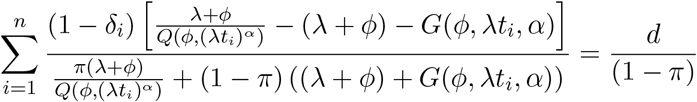

where 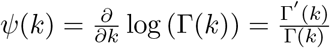 is the digamma function, 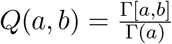 is the regularized gamma function; 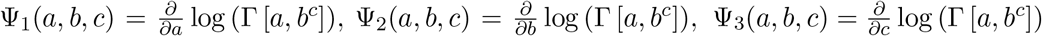 and 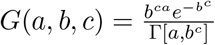 can be calculated numerically. Numerical methods need to be used to find the solution (maximum likelihood estimates) of these nonlinear equations.

Under mild regularity conditions, the MLEs are consistent, efficient and asymptotically normally distributed with a joint multivariate normal distribution given by

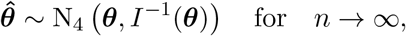

where *I*(*θ*) is the 4 × 4 Fisher information matrix for *θ*, and *I_ij_*(*θ*) is the (*i, j*)-th element of *I*(*θ*) given by

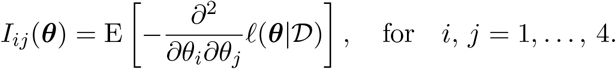

Note that it is not possible to compute the Fisher information matrix *I*(*θ*) due to the presence of censored observations (censoring is random and non-informative) and its complex form. Thus, one alternative approach is to use the observed information matrix *H*(*θ*) evaluated at 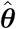, i.e. 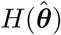, whose terms are given by

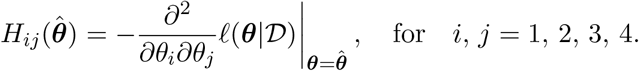

Large-sample (approximate) confidence intervals at level 100(1 − *ξ*)%, for each parameter *θ_i_*, *i* = 1, 2, 3, 4, can be calculated as

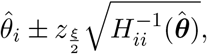

where *z*_ξ/2_ denotes the (*ξ/*2)-th quantile of a standard normal distribution.

## References

[1] Worldwide cancer statistics. https://www.cancerresearchuk.org/health-professional/cancer-statistics/worldwide-cancer”. Accessed: 2020-01-13.

[2] Araujo, L. H. d. L., C. S. Baldotto, M. Zukin, F. M. d. A. C. Vieira, A. P. Victorino, V. R. Rocha, R. C. Helal, J. H. Salem, N. Teich, and C. G. Ferreira (2014). Survival and prognostic factors in patients with non-small cell lung cancer treated in private health care. Revista Brasileira de Epidemiologia 17, 1001–1014.

[3] Berkson, J. and R. P. Gage (1952). Survival curve for cancer patients following treatment. Journal of the American Statistical Association 47 (259), 501–515.

[4] Brascia, D., G. De Iaco, M. Schiavone, T. Panza, F. Signore, A. Geronimo, D. Sampietro, M. Montrone, D. Galetta, and G. Marulli (2020). Resectable iiia-n2 non-small-cell lung cancer (nsclc): In search for the proper treatment. Cancers 12 (8), 2050.

[5] Chen, M.-H., J. G. Ibrahim, and D. Sinha (1999). A new Bayesian model for survival data with a surviving fraction. Journal of the American Statistical Association 94 (447), 909–919.

[6] Cline, M. S., B. Craft, T. Swatloski, M. Goldman, S. Ma, D. Haussler, and J. Zhu (2013). Exploring tcga pan-cancer data at the ucsc cancer genomics browser. Scientific reports 3, 2652.

[7] Cordeiro, G. M., V. G. Cancho, E. M. Ortega, and G. D. Barriga (2016). A model with long-term survivors: negative binomial birnbaum-saunders. Communications in Statistics-Theory and Methods 45 (5), 1370–1387.

[8] Ghitany, M., D. K. Al-Mutairi, N. Balakrishnan, and L. Al-Enezi (2013). Power lindley distribution and associated inference. Computational Statistics & Data Analysis 64, 20–33.

[9] Ghitany, M., F. Alqallaf, D. K. Al-Mutairi, and H. Husain (2011). A two-parameter weighted lindley distribution and its applications to survival data. Mathematics and Computers in simulation 81 (6), 1190–1201.

[10] Grambsch, P. M. and T. M. Therneau (1994). Proportional hazards tests and diagnostics based on weighted residuals. Biometrika 81 (3), 515–526.

[11] Liu, J., T. Lichtenberg, K. A. Hoadley, L. M. Poisson, A. J. Lazar, A. D. Cherniack, A. J. Kovatich, C. C. Benz, D. A. Levine, A. V. Lee, et al. (2018). An integrated tcga pan-cancer clinical data resource to drive high-quality survival outcome analytics. Cell 173 (2), 400–416.

[12] Louzada, F. and P. L. Ramos (2017). A new long-term survival distribution. Biostat Biometrics Open Acc J 1 (4), 1–6.

[13] Louzada, F., P. L. Ramos, and D. Nascimento (2018). The inverse nakagami-m distribution: A novel approach in reliability. IEEE Transactions on Reliability 67 (3), 1030–1042.

[14] Organization, W. H. et al. (2018). Latest global cancer data: Cancer burden rises to 18.1 million new cases and 9.6 million cancer deaths in 2018.

[15] R Core Team (2014). R: A Language and Environment for Statistical Computing. (Version 3.3.1). Vienna, Austria: R Foundation for Statistical Computing.

[16] Ramos, P., D. Nascimento, and F. Louzada (2017). The long term fréchet distribution: Estimation, properties and its application. Biom Biostat Int J 6 (3), 00170.

[17] Rodrigues, J., V. G. Cancho, M. de Castro, and F. Louzada-Neto (2009). On the unification of long-term survival models. Statistics & Probability Letters 79 (6), 753–759.

[18] Tammemagi, C. M., C. Neslund-Dudas, M. Simoff, and P. Kvale (2003). Impact of comorbidity on lung cancer survival. International journal of cancer 103 (6), 792–802.

[19] Xiong, J., Z. Bing, and S. Guo (2019). Observed survival interval: a supplement to tcga pan-cancer clinical data resource. Cancers 11 (3), 280.

